# The Anxiety and Pain of Fibromyalgia Patients during the COVID-19 Pandemic

**DOI:** 10.1101/2020.11.24.20188011

**Authors:** A. Y. Kharko, K. J. Hansford, P. L. Furlong, S. D. Hall, M. E. Roser

**Affiliations:** Faculty of Health, University of Plymouth; School of Psychology, Cardiff University; Faculty of Life & Health Sciences, Aston University

**Author notes:** **Correspondence:** Anna Y. Kharko, Lab 208, Link Building, University of Plymouth, Drake Circus, Devon PL4 8AA. Disclosures: Anna Kharko received funding from the School of Psychology, University of Plymouth. The grant provider was not involved in the conduct of the study. The authors have no conflicts of interest to declare.

**Keywords:** COVID-19, Pandemic, Anxiety, Fibromyalgia, Pain

## Abstract

**Background:** Early research on the impact of the COVID-19 pandemic found persistent related anxiety in the general population. We hypothesised that this anxiety will be associated with increased pain in chronic pain patients diagnosed with fibromyalgia (FM).

**Methods:** To study this, we carried out a 10-day online survey with 58 female participants, diagnosed with FM and no other pain condition. We identified which aspects of the COVID-19 pandemic evoked anxiety. We then asked participants to provide daily ratings of both anxiety and pain on 101-point visual analogue scales (VAS). Key participant characteristics were included as mediators in a mixed-effects analysis, where the primary outcome was pain VAS.

**Results:** We found that participants were most often anxious about *“impact on relationships”, “a family member contracting COVID-19”*, and *“financial hardships”*, but on average rated *“financial hardship”, “access to medication”*, and *“home loss/eviction”* as evoking the strongest anxiety. Mixed-effects modelling showed that an increase in pain was significantly associated with an increase in anxiety, when taking into account individual variance and daily caffeine intake. Age and intake of some mild analgesics were also linked to stronger pain.

**Conclusion:** Our results extend the initial findings from the literature about the effects of COVID-19 pandemic on chronic pain sufferers. We found that not only is pandemic anxiety in FM patients present, but it is associated with amplified self-assessed chronic pain.

**Significance:** The long-term support of fibromyalgia patients is challenging for healthcare professionals due to the nature of the condition. The new normal introduced by the pandemic particularly hinders pain management, which is the leading request from this patient group. Our study demonstrates that mental health decline during the COVID-19 pandemic is directly related to the worsening of pain in fibromyalgia. Core stressors that evoke the strongest anxiety were identified thus providing guidance for where to focus patient support.

## INTRODUCTION

In March 2020, Italy was the first European country to impose a nation-wide lockdown; restricting the behavioural freedom of its citizens in an attempt to control the spread of COVID-19. This step was soon followed by countries worldwide. The global consequences of the evolving circumstances were quickly apparent, with observed changes in behaviour, produced by contextual uncertainty (Shigemura et al., 2020). While the momentous impact of these changes have been widely posited, the negative impact of individual lived experiences on mental and physical wellbeing remains to be fully characterised.

Early research from China indicated significant decline in mental well-being, observed both in front-line healthcare workers (Li et al., 2020; Pappa et al., 2020) and the general population (Xiao et al., 2020). In the UK, when asked to describe their sentiment toward the pandemic, 55% of 2,500 pooled participants indicated high levels of anxiety (Kleinberg et al., 2020). The general consensus of the healthcare sector worldwide is that investigation of mental and physical well-being in the most vulnerable must follow. Here we propose to address this in a specific patient group, diagnosed with fibromyalgia (FM).

FM is a chronic musculoskeletal pain condition, accompanied by fatigue, nonrestorative sleep, and multiple comorbidities; common amongst which are anxiety disorders (Arnold et al., 2019). Generalised anxiety disorder alone is observed in 30% of FM sufferers (Gracely et al., 2012), but 60% exhibit symptoms consistent with diagnosis (Janssens et al., 2015). Despite the prevalence, anxiety in FM, is still poorly researched, compared to depression, which is indicated in 60% of patients (Walitt et al., 2015). This gap in literature is of particular importance since many patients believe distress is a key factor in the worsening of their pain (Bennett et al., 2007). Despite the increased base rate of anxiety, and patient reports of a causal link to pain, international guidelines make no specific recommendations on anxiety management in FM (Kia & Choy, 2017). The implications for the treatment gap that arise as a result, are likely to be compounded by the increase in the anxiogenic conditions during the current pandemic.

Recent research on chronic pain patients found an increase in pain during the pandemic; a rise attributed to lockdown and not the concurrently amplified anxiety (Fallon et al., 2020). These findings, however, are based on cross-sectional investigations that do not capture the high variability of anxiety and chronic pain. As recommended in a recent study, repeated assessment of both is necessary to study their relationship during the pandemic (Kleinberg et al., 2020).

To address these concerns, we conducted a 10-day survey of FM-diagnosed participants, with two aims. First, to determine which aspects of the pandemic were sources of anxiety in FM. Our next and primary aim was to determine the association between COVID-19 anxiety and FM pain.

We hypothesised that there will be several sources of anxiety for FM patients, each associated with different levels of anxiogenesis but contraction of the virus being the most prominent. We further hypothesised that an increase in COVID-19 anxiety will be associated with concurrently reported increase in pain.

## MATERIALS & METHODS

### Participants

The target population for the survey were adults, aged 18 to 60 years, diagnosed with FM for at least 6 months and no other pain condition. Participants were not recruited with COVID-19 relevant underlying health conditions, as reflected by placement on the NHS, UK Shielded Patient List. Participants were not recruited if taking gabapentinoids or hormone replacement therapy, both of which significantly mediate either pain or anxiety. Further screening was guided by the recently published diagnostic guidelines (Arnold et al., 2019). We included physical conditions that are commonly concurrent to fibromyalgia, if these were not associated with chronic pain. Mental health conditions classified under Axis I of Diagnostic and Statistical Manual of Mental Disorders, 5^th^ Edition (DSM-5) were also accepted.

### Screening Survey

To find eligible participants, a screening survey was launched online on Qualtrics (Qualtrics, Provo, UT). Its distribution began in May 2020 and involved circulation across various social media platforms and email newsletters of several FM charities and patient-lead support communities. The screening survey had the following sections.

### Participant Characteristics

Year of birth, gender, ethnicity, country of residence, marital status, weight, height, typical daily caffeine consumption and weekly alcohol consumption were recorded. Participants’ status as key workers or caregivers was recorded with details of circumstance (i.e. job or who they are caring for).

### Medical History & Medication

The Generalised Anxiety Disorder, Depression and PTSD sections from the Mini International Neuropsychiatric Interview (MINI) (Sheehan et al., 1998) were delivered online to account for the lack of a formal Axis I diagnosis in some participants. A researcher reviewed the answers to calculate scores for the PTSD, GAD, and depression, as most relevant to the study sections. As part of medical history, participants confirmed details of their FM diagnosis (e.g. year, diagnosis route). Participants also answered a series of questions to eliminate the possibility of excluding secondary circumstances (ESCs) that may indicate an undiagnosed chronic condition (e.g. convulsions/seizures). Based on COVID-19 advice from the NHS at the time of screening pregnancy was also considered ESC.

Medication intake in the last 30 days was recorded to identify participants, who qualify as a shielded person but failed to select a relevant option (e.g. asthma inhaler).

### Experience with Pandemic & Anxiety

Participants were screened for possible COVID-19, based upon the cardinal symptoms, identified by the NHS. Any participant reporting positively were excluded from participation.

Participants were then asked whether they had experienced anxiety that is related to the COVID-19 pandemic. Respondents were to select as many as applicable from 14 possible categories (see Supporting Information 1). The categories were predetermined by the research team based on recent publications and conversations with stakeholders). Any category that was not selected was to be removed from the daily surveys. All categories were selected; thus, none were discarded.

### First Daily Survey

#### Pain History & General Anxiety

The first daily survey acquired baseline measures of participant’s recent pain history and dispositional anxiety. The presence of FM symptoms was reconfirmed though the American College of Rheumatology Criteria, ACR, (Wolfe et al., 2010, 2016). Impact of FM on daily functioning was measured through the Revised Fibromyalgia Impact Questionnaire, FIQ-R (Bennett et al., 2009), and on overall pain through the Short Form McGill Pain Questionnaire, MPQ-SF (Melzack, 1987). To assess dispositional anxiety, we used the trait version of the State-Trait Inventory for Cognitive and Somatic Anxiety, STICSA-T (Grös et al., 2007).

#### Daily Caffeine, Alcohol, & Medication

Several mediators of pain and anxiety were expected. First is caffeine, which may perpetuate anxiety (Nardi et al., 2009), but is also considered an adjuvant analgesic (Sawynok, 2011). Alcohol is also a known pain mediator. A curvilinear relationship is observed between chronic pain and the consumption of alcohol (see (Zale et al., 2015) for a review). Lastly, our target population was expected to take a variety of medication. To measure, and ultimately control for, these variables, we recorded daily the intake of caffeine (in mg), alcohol (in units), and medication (see Analysis).

#### Daily Anxiety & Pain

Daily ratings of anxiety and pain were the primary variables. To measure anxiety, participants first identified the pandemic-related sources of anxiety relevant to that day. Only then they provided anxiety ratings for the selected sources on a 101-point visual analogue scale (aVAS). If an anxiety source was skipped, that source was automatically recorded as ‘0’ – no anxiety. Pain was similarly recorded on a 101-point visual analogue scale (pVAS). For both scales ‘0’ represented no anxiety/pain, ‘1’ was minimal anxiety/pain and ‘100’ was the worst possible anxiety/pain.

### Daily Survey 2 – 10

The same surveys were completed for 9 more consecutive days, omitting the pain history and general anxiety questionnaires.

### Procedure

The study was carried out in May and June 2020. Eligible participants completed the first of the 10-days survey within two weeks of passing screening. This resulted in multiple waves of participants with various start times. The daily survey was circulated at 4 PM local time to the participant, followed by a reminder at 8 PM and 10 AM the next day. Participants were given up to 24 hrs after the initial email for survey completion. Answers beyond the time limit were considered as missed. Participants were allowed to miss up to two surveys to achieve a sample completion rate no lower than 80%.

The study was granted ethical permission by the ethical board in the Faculty of Health at the University of Plymouth and conformed to the Declaration of Helsinki. All data was stored anonymously. Informed consent was gathered during screening and each daily survey. Debrief was provided upon study completion. The data that support the findings of this study are available from the corresponding author upon reasonable request.

### Analysis

Following our aims, we first established whether FM patients experienced anxiety related to the pandemic by asking participants to select and rate applicable sources of anxiety on each daily survey. Mean (μ) and standard deviation (*SD*) were calculated for the provided ratings, ignoring those that were automatically zeroed.

We then investigated the relationship of the experienced COVID-19 anxiety with FM pain. Experienced COVID-19 anxiety was defined as the daily average anxiety rating, or aVASμ. It was calculated using only the provided anxiety ratings from the selected anxiety sources. Sources that were not selected by the participant as anxious received an automatic rating of 0 and were thus not contributing aVASμ. This focused approach allowed us to align anxiety ratings between participants with considerably different numbers of selected sources. More importantly, it is the aggregate score that encapsulates COVID-19 pandemic anxiety as a whole and not the separate categories, which capture only individual aspects of that anxiety. FM pain was defined directly through the pVAS rating.

We hypothesised that pVAS would increase with the increase of aVASμ. However, we also expected confounding factors so we utilised mixed-effects modelling. pVAS was the outcome variable. To determine a model of best fit, the following covariates were possible: aVASμ, participant (*Participant*), participant’s age (*Age*), participant status as key worker (*Key Worker*) or caregiver (*Caregiver)*, daily caffeine (*Caffeine*), and daily alcohol (*Alcohol*). We also created a categorical variable that denotes the presence of a concurrent medical diagnosis (*Comorbidity)*. It was data-driven and resulted in the following levels: no comorbidity, anxiety/fear disorder, depression, allergy, and thyroid condition. Daily medication vastly varied between participants to be added as a single covariate. We thus created separate categorical covariates for medication that was present in at least 10% of the total number of daily surveys (excluding supplements and nutraceuticals). The following covariates emerged: over the counter analgesics (*OTCs)*, non-steroidal anti-inflammatory drugs (*NSAIDs*), opioids (*Opioids*), tricyclic antidepressants (TCAs), and SSRIs (*SSRIs*). Date of survey completion was not used as a factor in analysis since data was not balanced in respect to calendar date.

Analysis was carried out in R Studio (v 1.2.5). Power analysis was carried through simulations (*n* = 200) in simr (v 0.4), where Kenward-Roger approximation was calculated for each fixed covariate to reach effect size *d* = .3 at ≥ 80% (Green & MacLeod, 2016; Matuschek et al., 2017). A Kenward-Roger approximation was calculated for each fixed covariate. For hypothesis testing, models were compared through likelihood-ratio testing and Bayes factor against a baseline model consisting of participant-only intercept. The best model was chosen based on fit assessed through Bayesian modelling, following analysis protocol described elsewhere (Muth et al., 2018), as well as BIC and AIC. Finally, the predictions of the best fit model generated by lme4 (v 1.1.21) were confirmed through Bayesian credible intervals from rstanarm (v 2.18.2).

## RESULTS

Out of 455 screened participants, 397 (87%) were excluded due to various reasons (see Fig. 1). Of those who qualified, 58 completed ≥ 80% of the surveys and their data were analysed.

**Figure 1.**
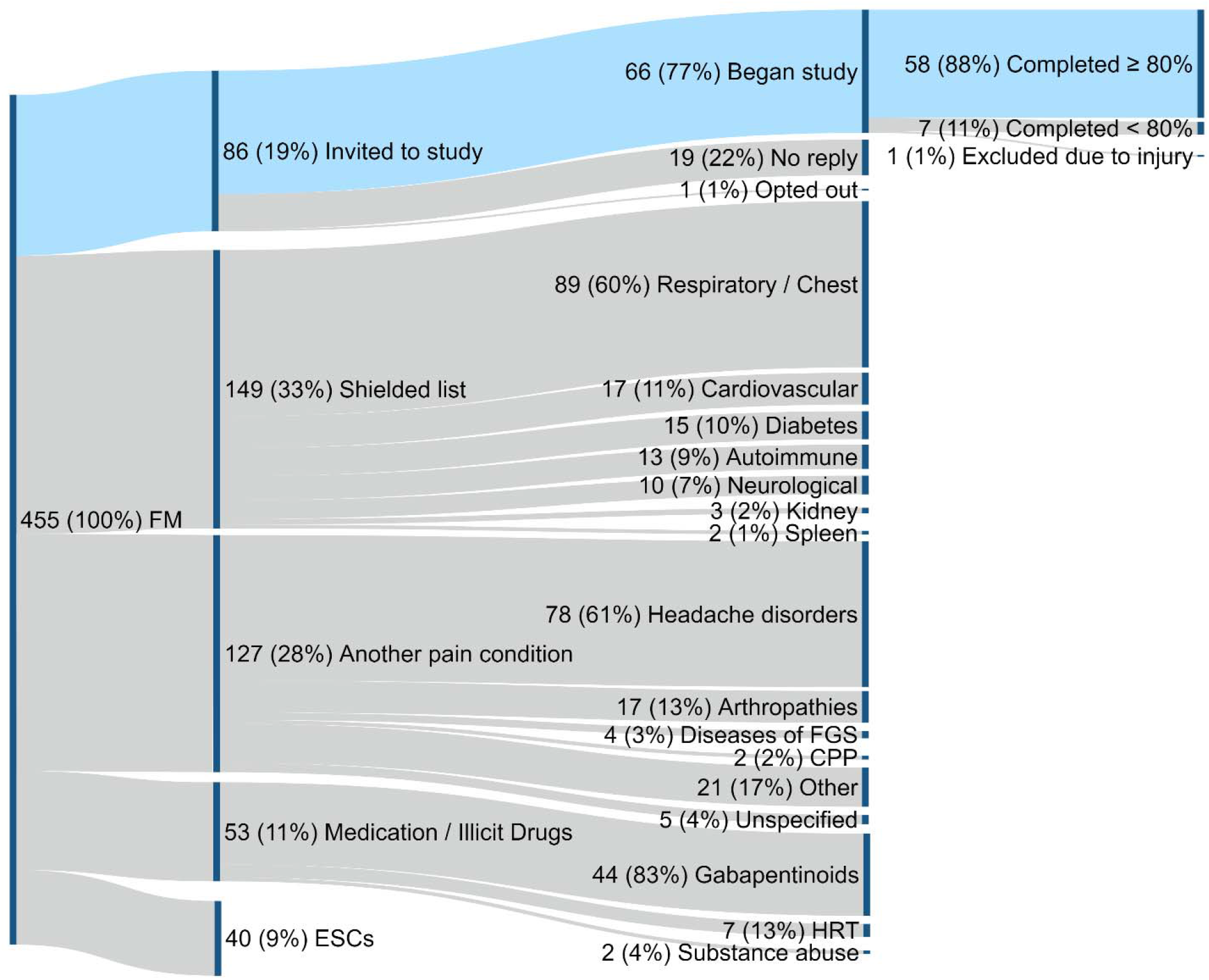
Participant recruitment diagram. Participants were excluded based on the following criteria (in order): 1) presence of fibromyalgia (FM) diagnosis; 2) presence of conditions from the shielded list; 3) presence of another pain condition, e.g. diseases of the female genital system (FGS) or chronic primary pain condition (CPP); 4) intake of medication such as gabapentinoids, hormone replacement therapy (HRT), or illicit drugs; and lastly 5) presence of excluding secondary circumstances (ECSs). Each node is labelled with number of participants (n), percent from total screened sample (%) and reason for inclusion/exclusion.

Participant characteristics can be found in Table 1.

**Table 1.** Sample Characteristics.

| | $\mu$ or n | (SD) or % |
| --- | --- | --- |
| Gender (female) <sup>a</sup> | 58 | 100% |
| Age (n years) | 40.29 | (10.98) |
| Ethnicity <sup>a</sup> |  |  |
| *Black / African / Caribbean* | 1 | 2% |
| *Caucasian* | 56 | 96% |
| *Hispanic* | 1 | 2% |
| Weight (kg) | 82.92 | (23.67) |
| Height (cm) | 166 | (0.7) |
| BMI | 30.16 | (8.07) |
| Country of Residence <sup>a</sup> |  |  |
| *Canada* | 1 | 2% |
| *Spain* | 1 | 2% |
| *UK* | 52 | 89% |
| *USA* | 4 | 7% |
| Marital Status <sup>a</sup> |  |  |
| *Civil Partnership / Married* | 26 | 45% |
| *Separated / Divorced* | 10 | 17% |
| *Single* | 20 | 35% |
| *Widowed* | 2 | 3% |
| Key worker (n yes) <sup>a</sup> | 22 | 38% |
| Caregiver (n yes) <sup>a</sup> | 16 | 28% |
| Fibromyalgia (ICD-11 MG30.01) |  |  |
| *Diagnosis duration (years)* | 3.86 | (3.79) |
| *Symptoms duration (years)* | 9.15 | (7.37) |

$\mu$ - average value, n – count, SD – standard deviation, % - percentage.
<sup>a</sup> Items, for which n and % were calculated.
<sup>b</sup> Medication of various categories that are not relevant to the study objectives (e.g. contraception, topical creams, digestion medication).

Comorbidities, medication intake, and questionnaire scores of the final sample are summarised in Table 2.

**Table 2.** Medical Comorbidities, Medication Intake and Questionnaires.

|  | **μ or n** | **(SD) or %** |
| --- | --- | --- |
| Medical Comorbidities (present in n participants) <sup>a</sup> | 28 | 48% |
| *Unspecified Allergy (ICD-11 n)* | 7 | 12% |
| *Allergic rhinitis due to pollen (ICD-11 CA08.00)* | 5 | 8% |
| *Anxiety/fear-related disorder (ICD-11 6B00 – 6B0Y)* | 2 | 3% |
| *Obsessive-compulsive disorder (ICD-11 6B20)* | 1 | 2% |
| *Panic attacks (ICD-11 MB23.H)* | 10 | 17% |
| *Depressive disorder (ICD-11 6A71)* | 2 | 3% |
| *Hypothyroidism (ICD-11 5A00)* | 5 | 8% |
| *Postviral fatigue syndrome (ICD-11 8E49)* | 4 | 7% |
| *Raynaud phenomenon (ICD-11 BD42)* | 1 | 2% |
| *White coat hypertension (ICD-11 MC80.00)* | 1 | 2% |
| Medication (taken by n participants in the last month) <sup>a</sup> | 54 | 93% |
| *Analgesics (non-prescribed)* | 21 | 36% |
| *Beta-blockers* | 2 | 3% |
| *Antihistamines* | 10 | 17% |
| *Benzodiazepines* | 1 | 2% |
| *IBS Medication / PPIs* | 8 | 14% |
| *Muscle relaxants* | 2 | 3% |
| *NSAIDs* | 20 | 35% |
| *Opioids* | 22 | 38% |
| *TCAs* | 13 | 22% |
| *Thyroid Medication* | 6 | 10% |
| *SNRIs* | 4 | 7% |
| *SSRIs* | 12 | 20% |
| *Supplements / Nutraceuticals* | 19 | 33% |
| *Other* <sup>b</sup> | 15 | 26% |
| ACR (n met criteria) | 58 | 100% |
| FIQ-R | 57.58 | (16.95) |
| SF-MPQ |  |  |
| *Pain Descriptors (0 - 45)* | 20.36 | (7.09) |
| *Sensory* | 15.74 | (5.65) |
| *Affective* | 4.62 | (2.31) |
| *Visual Analogue Scale (0 - 10)* | 6.39 | (1.72) |
| *Present Pain Intensity (0 - 5)* | 3.03 | (.65) |
| Trait STICSA | 48.76 | (12.7) |
| *Cognitive Anxiety* | 24.19 | (7.32) |
| *Somatic Anxiety* | 24.57 | (6.64) |
| M.I.N.I.<sup>a</sup> |  |  |
| *Depression (n meet criteria)* | 33 | 57% |
| *PTSD (n meet criteria)* | 24 | 41% |
| *Anxiety (n meet criteria)* | 51 | 88% |

*IBS* – irritable bowel syndrome; *PPIs* – proton-pump inhibitors; *NSAIDs* - non-steroidal anti-inflammatory drugs; *TCAs* – tricyclic antidepressants; *SNRIs* – serotonin–norepinephrine reuptake inhibitors; *SSRIs* – selective serotonin reuptake inhibitors; *ACR* – American College of Rheumatology Criteria for Fibromyalgia; *FIQ-R* – Revised Fibromyalgia Impact Questionnaire; *SF-MPQ* – Short-form McGill Pain Questionnaire; *STICSA* – State-Trait Inventory for Cognitive and Somatic Anxiety; *M.I.N.I.* – Mini-International Neuropsychiatric Interview; *PTSD* – post-traumatic stress disorder.
μ - average value, n – count, *SD* – standard deviation, % - percentage.
<sup>a</sup> Items, for which n and % were calculated.
<sup>b</sup> Medication of various categories that are not relevant to the study objectives (e.g. contraception, topical creams, digestion medication).

Due to 4 missed daily surveys, the final dataset comprised of 576 daily surveys. In total participants made 1,600 ratings across all anxiety sources. However, the anxiety source ‘*other’*, which contained participant-entered reasons was highly heterogeneous and commonly unrelated to the pandemic. For this reason, it was removed from analysis leaving 1,548 anxiety ratings. On average participants chose 2.79 (*SD* = 2.27) sources per day out of the remaining 13. Fig. 2 shows the anxiety sources ordered by mean rating in a descending order, with the researcher-calculated cumulative rating aVASμ on top.

**Figure 2.**
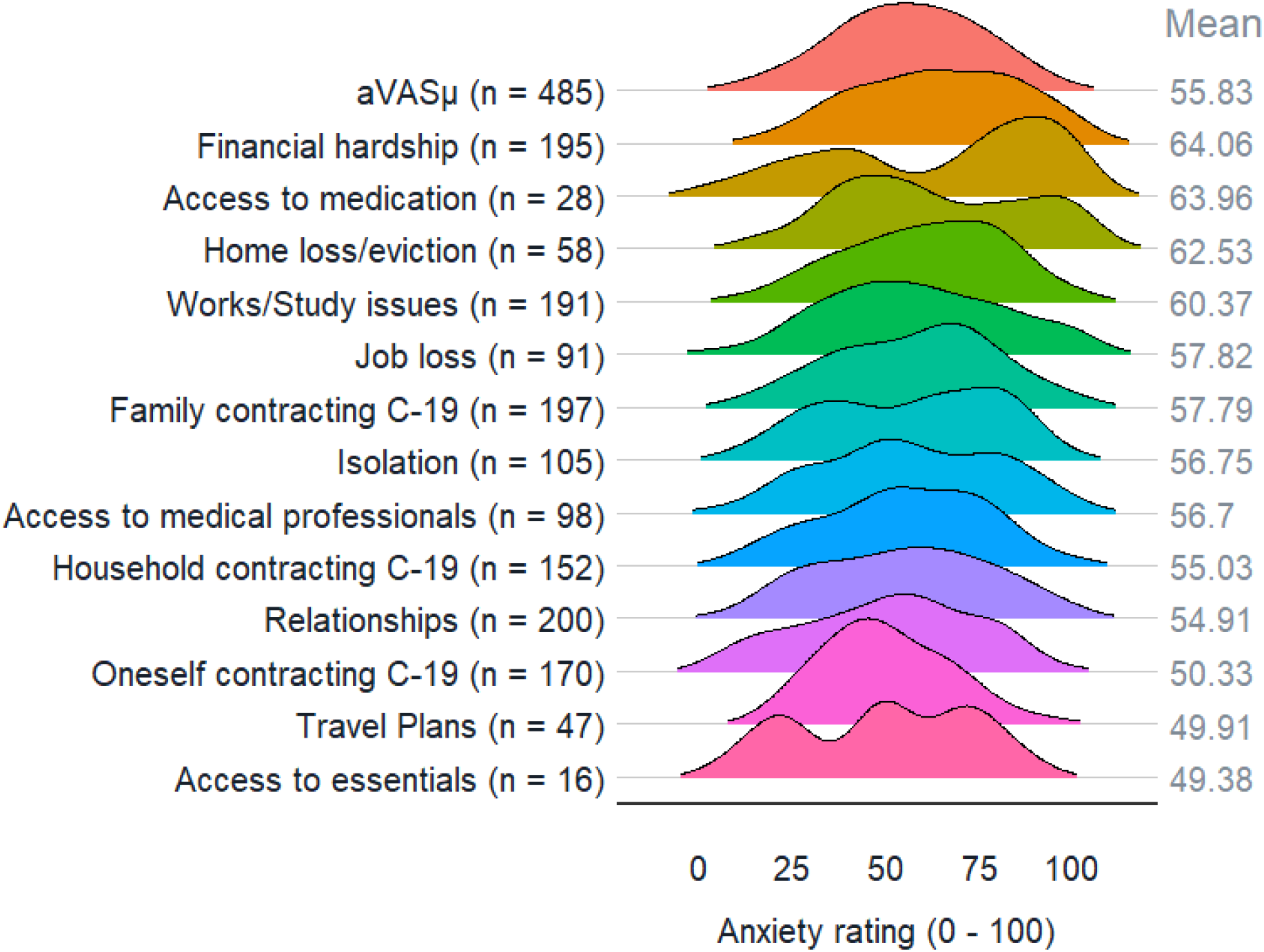
Distributions of anxiety ratings per source. The first row of the half-violin plot shows the distribution of the daily average anxiety ratings (aVASμ). Subsequent rows show the distribution of ratings per anxiety source, ordered by highest rated on average to the lowest. The number of times an anxiety source was selected (across all daily surveys) is shown in brackets. The average anxiety rating per source is shown on the right of each distribution.

After we determined the different sources of anxiety related to the pandemic, we analysed the relationship between experienced anxiety and FM pain. Plotting the anxiety and pain ratings on a scatterplot (see Figure 3) suggests that a positive trend does exist between the two.

**Figure 3.**
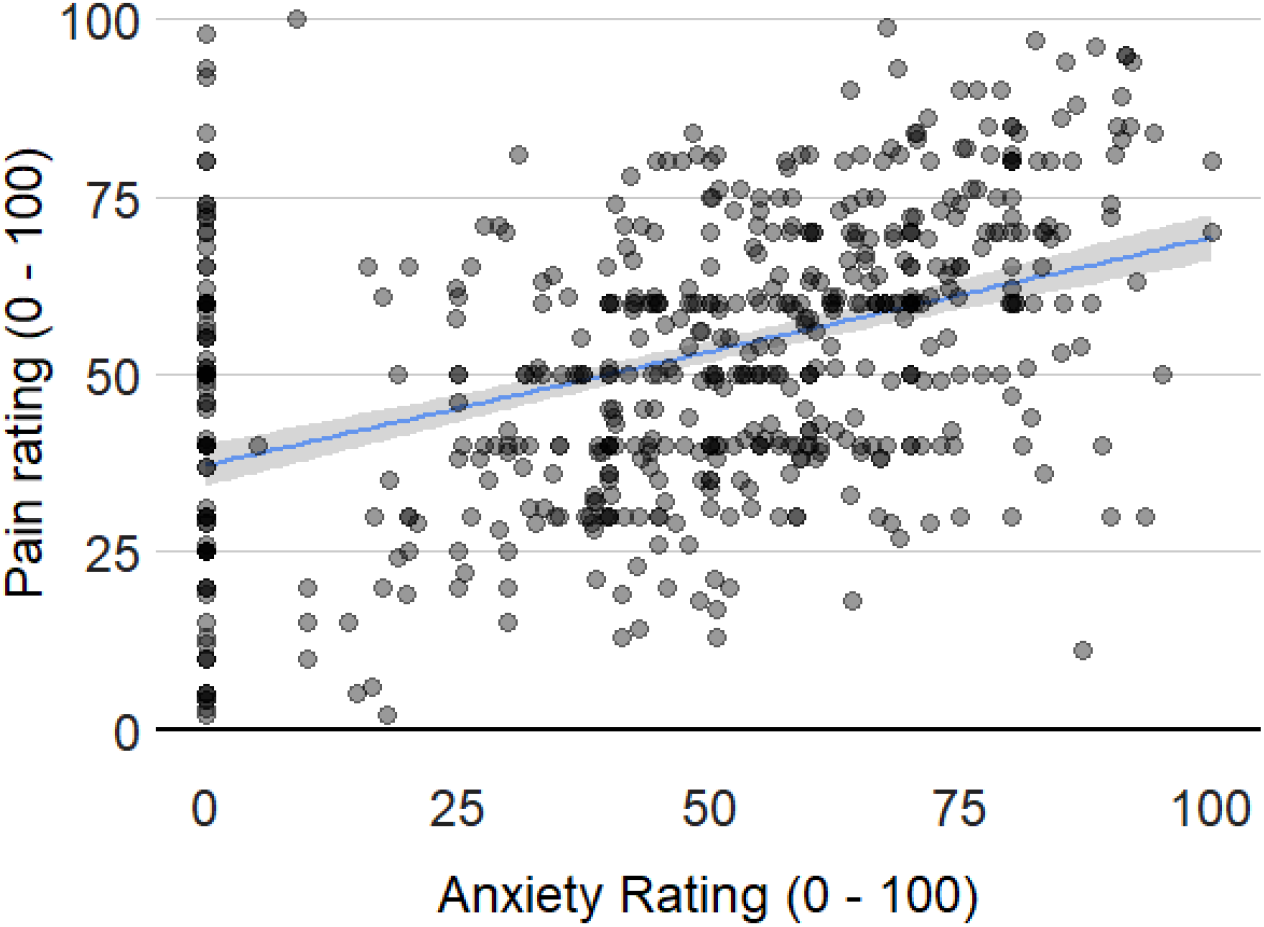
Scatterplot of anxiety and pain ratings. The daily anxiety rating (aVASμ) is plotted on the x-axis and the daily rating of fibromyalgia pain (pVAS) is on the y-axis. The regression line is a function of x ∼ y, the shaded area represents the standard error around it.

Mixed-effects modelling showed that the best fit model had the following formula:

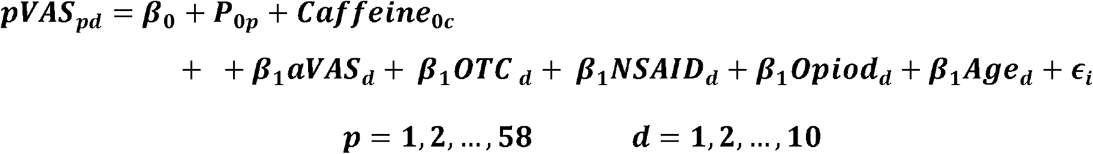

The best fit model significantly differed from the baseline model, χ^2^ = 94.16, *p* <.001, BF = 1.04 x10^14^. It contained several covariates that were significantly linked to pain (see Table 3). As predicted, the model showed that higher self-assessed FM pain (pVAS) was significantly associated with increased daily anxiety (aVASμ). Further, respondents who took OTCs and NSAIDs reported more pain than those who did not. This difference was mirrored in patients who took opioids but the association failed to reach significance. Intake of TCAs was not part of the best fit model, possibly due to the lower number of observations. OTC analgesics (17%), NSAIDs (17%), and opioids (22%) accounted for a large portion of total medication intake, while TCAs were present in only 10% of the measurements. In contrast, SSRIs were common (21%) but still were not part of the best fit model.

**Table 3.** Best Fit Model Statistics.

|  | Traditional Analysis |  |  | Bayesian Analysis |  |  |  |
| --- | --- | --- | --- | --- | --- | --- | --- |
|  | Coeff. | (SE) | t-value | Coeff. | (SD) | CI |  |
|  |  |  |  |  |  | 2.5% | 97.5% |
| Fixed effects |  |  |  |  |  |  |  |
| Intercept | 18.74 | (5.87) | 3.19 ** | 18.65 | (5.95) | 7.34 | 30.54 |
| aVAS | .21 | (.03) | 7.27 *** | .2 | (.03) | .15 | .26 |
| OTCs Intake | 7.7 | (2.18) | 3.53 *** | 7.71 | (2.23) | 3.43 | 12.07 |
| NSAIDs Intake | 8.01 | (2.16) | 3.71 *** | 7.94 | (2.2) | 3.57 | 12.29 |
| Opioids Intake | 4.4 | (2.32) | 1.9 † | 4.39 | (2.36) | -.32 | 8.96 |
| Age | .5 | (.14) | 3.68 *** | .5 | (.14) | .23 | .78 |
| | $\sigma^2$ | (SD) | | | | | |
| Random effects |  |  |  |  |  |  |  |
| Participant | 1.1 | (10.49) |  |  |  |  |  |
| Caffeine Intake (mg) | 1.49 | (.01) |  |  |  |  |  |
| Residual | 1.82 | (13.50) |  |  |  |  |  |

In frequentist analysis, the linear mixed model was fit by maximum likelihood and t-tests were calculated automatically using Satterthwaite approximations to degrees of freedom. In Bayesian analysis, weakly informed priors were used to estimate parameters.
aVAS – anxiety visual analogue scale rating, OTCs – over the counter analgesics, NSAIDs – non-steroid anti-inflammatory drugs, *Coeff.* – predicted value for coefficient, $\sigma^2$ – estimated variance, *SE/SD* – estimated standard error/deviation, *CI* – credible interval.
\*\*\* $p < .001$ , \*\* $p < .01$ , \* $p < .05$ , <sup>a</sup> $p = .058$

Age was also significantly allied to pain: the older a participant was, the higher their reported pain was. Variance in pain was attributed to individual differences between participants and the daily amount of caffeine. Models including presence of comorbid conditions and daily alcohol intake were not superior.

## DISCUSSION

The present study investigated the effects of the COVID-19 pandemic on the well-being of FM patients. We determined which aspects of living during the pandemic are anxiogenic to FM patients and assessed daily fluctuation in self-reported pain. We found that although respondents differed in sources of anxiety day-to-day and between each other, the resulting incline in anxiety ratings was reflected in ratings of chronic pain.

On the basis of discussions with stakeholders, we expected, that as a health-compromised group, our respondents would be most concerned with health-related issues: *“oneself contracting COVID-19”, “access to medication”*, and *“access to medical professionals”*. Instead, our findings on anxiety sources agree with recently published surveys of the general population (Kleinberg et al., 2020; Shahabi et al., 2020). On average FM patients rated *“financial hardship”, “access to medication”*, and *“home loss/eviction”* as the most anxiogenic. Similar concerns with economic security have been observed in both vulnerable and general populations (Kleinberg et al., 2020; McElroy et al., 2020). In that, FM respondents appear homogenous with their healthy counterparts. However, FM patients were least anxious about *“oneself contracting COVID-19”, “delayed travel plans”* and *“access to essentials”*. The former has been reported as both the most prevalent and the strongest anxiety source (Kleinberg et al., 2020). It is unclear as to why FM respondents rated disease contraction so low. It is possible that as self-identified vulnerable group participants took measures they perceived to sufficiently reduce their risk of contracting COVID-19. Importantly, the highest rated anxiety sources were not necessarily the most reported.

Most often, FM patients pointed to *“impact on relationships”, “family contracting COVID-19”*, and *“financial hardship”* as evoking anxiety. Fear over potentially negative effects of long-term quarantine on interpersonal relationships has been predicted in the literature (Schimmenti & Starcevic, 2020), as has concern over family members contraction risk rather than oneself (Wang et al., 2020). The high prevalence of financial anxiety fortifies it as a central concern for FM patients. Economic downfall is expected to maintain for years to come (Baldwin, 2020), which raises the question whether it will remain anxiogenic for chronic pain patients. Pre-pandemic up to 46% of FM sufferers reported job loss due to health complications (Al-Allaf, 2007). Redundancy during the pandemic may mean that such individuals will struggle with finding new employment due to the work restrictions imposed by FM (Bossema et al., 2012). Further, worsening of socio-economic status will likely put FM individuals in a higher risk for contracting COVID-19 (Patel et al., 2020), thus this group of patients should become focus of attention for policymakers. In contrast, FM patients pointed the least times to “*delayed/cancelled travel plans”, “access to medication”*, and *“access to essentials”* as anxiogenic. Even though this is also in agreement with cited research, our findings cover only the early phase of the COVID-19 pandemic (i.e. the “first wave”). Easing of travel restrictions may introduce new stressors as it was seen in Wuhan, China (Ma et al., 2020). At present, we found that participants who utilised the comments box at the end of the daily surveys welcomed some easing of the travel rules:

> *“… The PMs [Prime Minister’s] update has been great as it means we could have some family over and it looks like our caravan holiday in August will go ahead, finally something to look forward to*.*”*

An in-depth qualitative analysis of these comments is a topic of another publication (in preparation). We did observe a reduction in some anxiety ratings coinciding with easing of restrictions in the UK (see Supporting Information 2).

The central finding that emerged during analysis was the positive relationship between anxiety and pain ratings. Our model predicts that for each 10 points of increase on aVAS scale, FM respondents will experience a 2-point increase in pVAS. Although this increase is small, particularly when compared to the increase associated with intake of OTC and NSAIDs, it is significant. While FM patients experience psychosomatic symptoms, the pain chronicity is not given rise to by a psychogenic mechanism (Pikoff, 2010). Thus, daily psychological fluctuations cannot be presumed as leading determinant of FM pain. For as long as the origin of this pain remains unknown, factors mediating it are actively sought by patients and researchers. The increase of pVAS ratings observed during OTC and NSAID analgesics is indicative of patient behaviour. Both are part of self-management plan for many FM patients despite inconclusive clinically significant benefits (Derry et al., 2017; Rocha et al., 2020). We therefore interpret our findings as indicative of intake of these analgesics in the presence of heightened pain.

Another important finding is that apart from age, no other participant characteristic was found to significantly improve the best fit regression model. A plausible explanation for that is the scope of measured participant qualities was not sufficient. Presence of mental distress or a clinical diagnosis of anxiety have been linked to poor quality of life and pain in FM, unlike comparable pain conditions (Gormsen et al., 2010). In our sample, only 3% participants had an anxiety as a diagnosis but 88% had an indication of general anxiety as measured through MINI.

The finding that the pandemic is not only conducive to heightened anxiety but that the resulting emotional distress is reflected in FM pain, has several implications. Foremost, it confirms the previously raised concerns that the “new normal” introduced by the pandemic may qualitatively differently impact vulnerable populations. Second, it highlights the relationship between mental and physical wellbeing in FM pain. Evidence-based international guidelines suggest that treatments target foremost patient-reported complaints (Clauw, 2014; Häuser et al., 2010). Psychiatric evaluation is instead undertaken upon request. This practice is in contrast with research where presence of psychological distress has been linked to disease progression (Marcus, 2009). Our findings support that literature and evidence the necessity to address anxiety in FM during the pandemic. Cognitive and behavioural interventions, however, are often dismissed by FM patients as indicated by low treatment adherence (Dobkin et al., 2006). Further, such therapies may not be viable during a pandemic. New alternative options are needed to aid mental health in patients for the benefit of their chronic pain. One promising direction are open-label placebos, which combine the traditional administration of a placebo pill with an informative narrative (Locher et al., 2017). Such placebos have been found to improve psychological symptoms and with them chronic physical complaints (Carvalho et al., 2016; Kelley et al., 2012; Sandler & Bodfish, 2008).

Lastly, we identified several limitations of our study. The primary focus of our research was the experience of pain in the presence of COVID-19 anxiety. Pain, however, is only one of the cardinal FM symptoms. Sleep disturbances and fatigue are both most reported concerns for FM (Arnold et al., 2008), as well as both are acutely sensitive to stressors (Affleck et al., 1996). There has been a decline in the quality of sleep in the general population (Casagrande et al., 2020). If the regression is similar in FM, it may have profound consequences for pain (Affleck et al., 1996). Future investigations should therefore integrate daily evaluations of fatigue and sleep alongside of assessments of COVID-19 anxiety. Another constraint of our work was that the survey was carried out during the early phase of the pandemic, when anxiety was at its highest. Observing pain during time of relative normality as well as during reverting of lockdown would be advantageous for the understanding of long-term development of COVID-19 anxiety in the presence of chronic pain.

## CONCLUSION

Our study found that FM pain increased with COVID-19 anxiety during the pandemic. This relationship was mediated by individual differences and intake of certain medication, such as OTC analgesics and NSAIDs, together with caffeine consumption. These findings indicate that mental health decline as a result of the COVID-19 pandemic coincides with worsening of the physical wellbeing in chronic pain sufferers. Further research is necessary to broaden the understanding of how other key FM symptoms, fatigue and sleep disturbances, are impacted during the pandemic.

## Data Availability

Data is available upon request to the corresponding author.

## Acknowledgements

The authors thank Carol Hambly, the organiser of a Fibromyalgia Support Group at Plymouth, UK, for her continued support for fibromyalgia research by providing invaluable insight and feedback. The authors also thank Fibromyalgia Action UK, Fibromyalgia Awareness UK, and Pain UK for their assistance with recruitment.

## Author Contributions

AK, KH, PF, SH and MR were responsible for the conceptual discussions, design and data interpretation. AK and KH collected and analysed data. All authors contributed to the critical revision of the manuscript and provided approval of the final version.

