## Supporting Information 1 for "The Anxiety and Pain of Fibromyalgia Patients during the COVID-19 Pandemic"

**Anxiety Sources**

1) Anxiety about oneself contracting COVID-19 coronavirus;

2) Anxiety about a household member contracting COVID-19 coronavirus;

3) Anxiety about a family member contracting COVID-19 coronavirus;

4) Anxiety about work/study issues due to the pandemic;

5) Anxiety about job loss due to the pandemic;

6) Anxiety about financial hardships due to the pandemic;

7) Anxiety about home loss/eviction due to the pandemic;

8) Anxiety about access to medication during the pandemic;

9) Anxiety about access to medical professionals during the pandemic;

10) Anxiety about access to essential supplies during the pandemic;

11) Anxiety about isolation during the pandemic;

12) Anxiety about delayed/cancelled travel plans due to the pandemic;

13) Anxiety about impact on personal relationships (friendships, romantic, family, or other);

14) Anxiety about participant-entered reason due to the pandemic. ^†^

^†^ Not included in the analysis due to the variability of answers, many of which not related to the pandemic.
