## Supporting Information 2 for "The Anxiety and Pain of Fibromyalgia Patients during the COVID-19 Pandemic"

**Daily Changes in Anxiety Ratings Categorised by Source**

Ratings were provided by fibromyalgia-diagnosed participants. Similarly to the analysis, data includes only ratings above 0. Where applicable, the authors have marked significant events which coincided with large trend change.

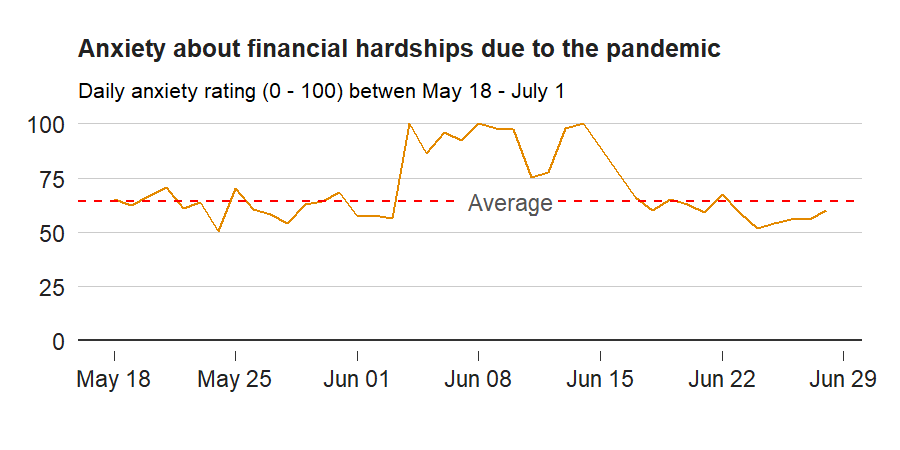

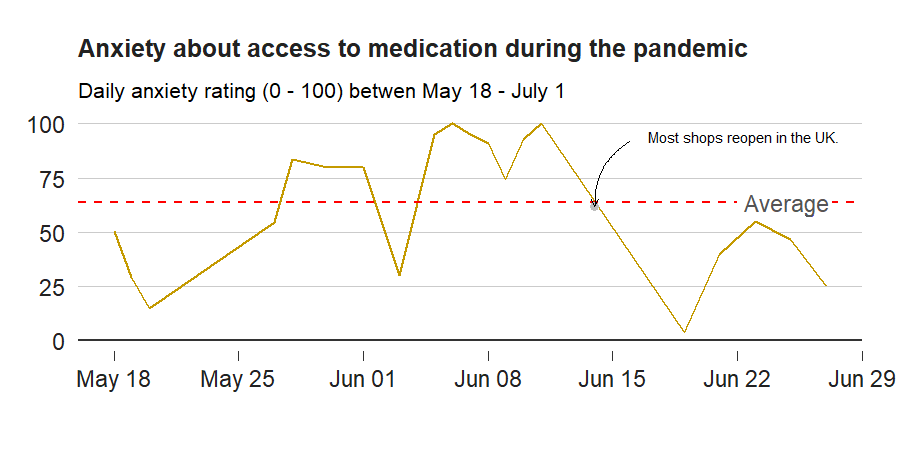

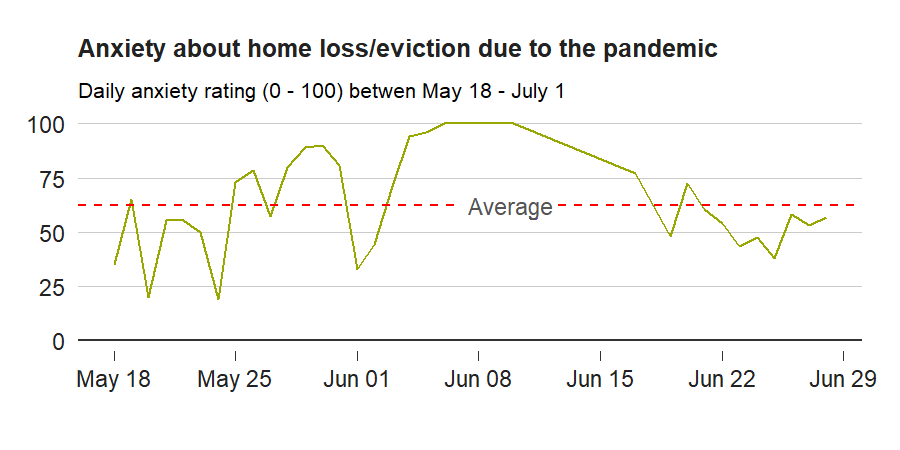

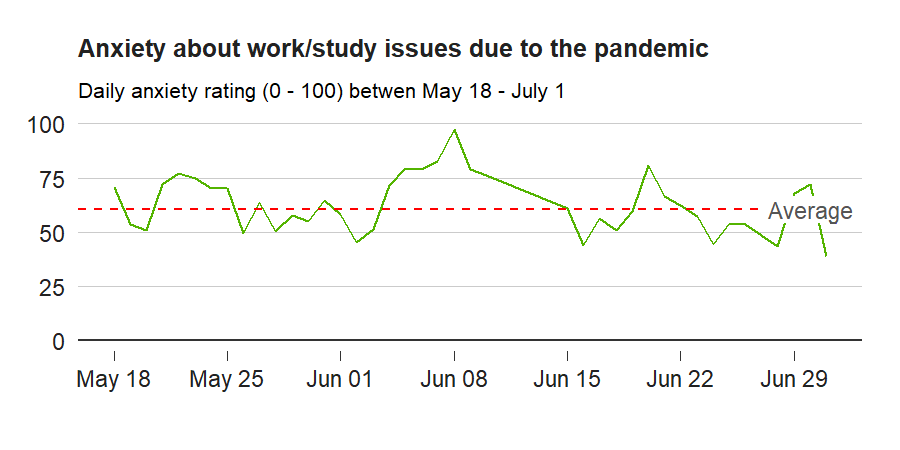

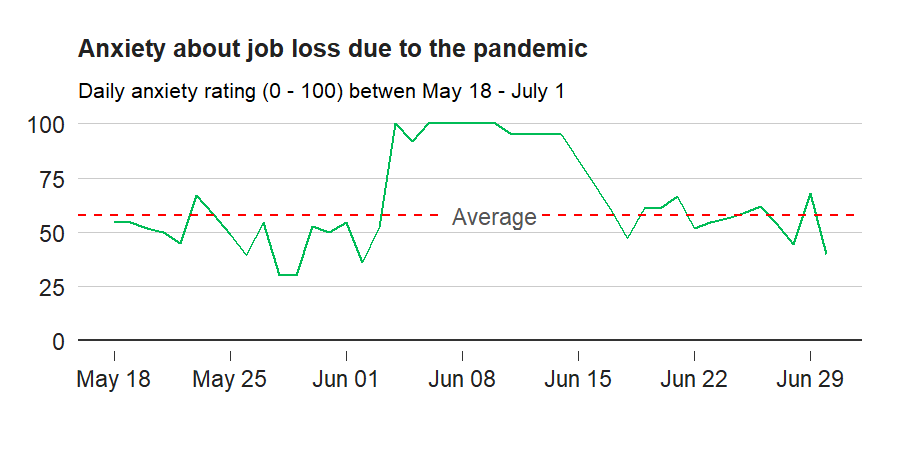

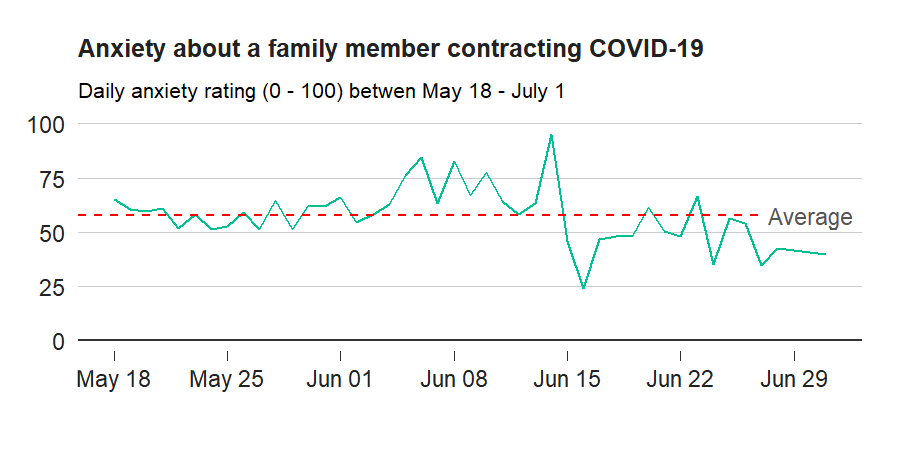

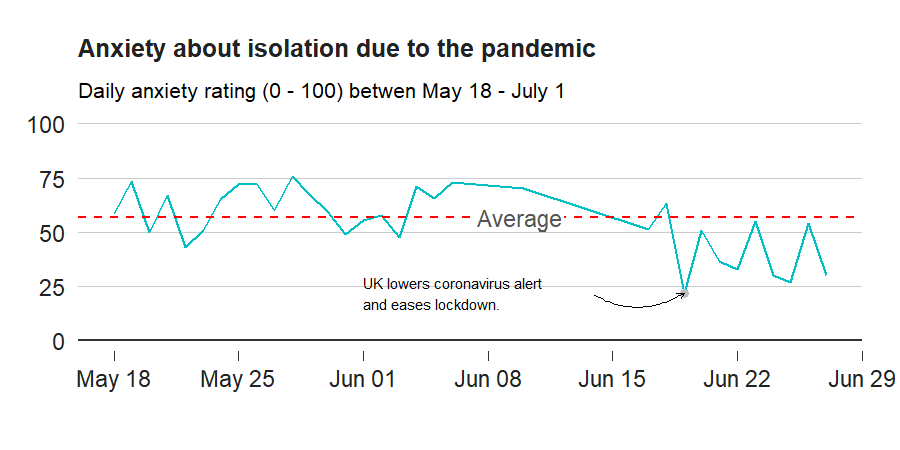

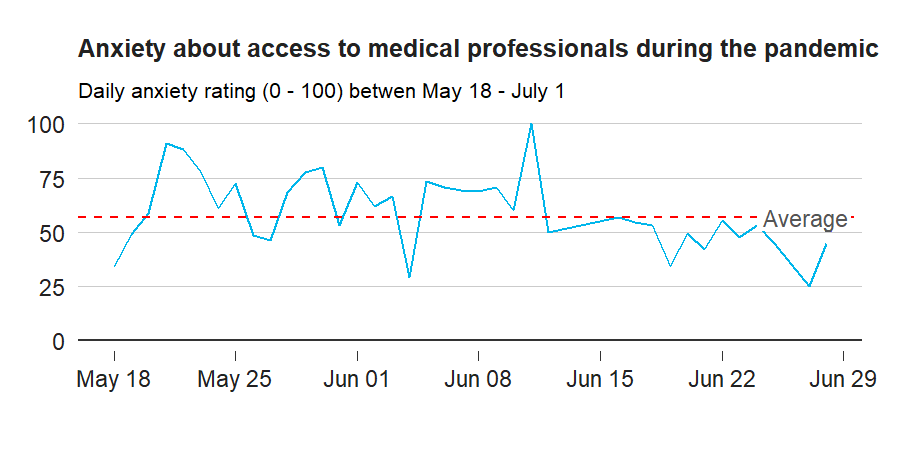

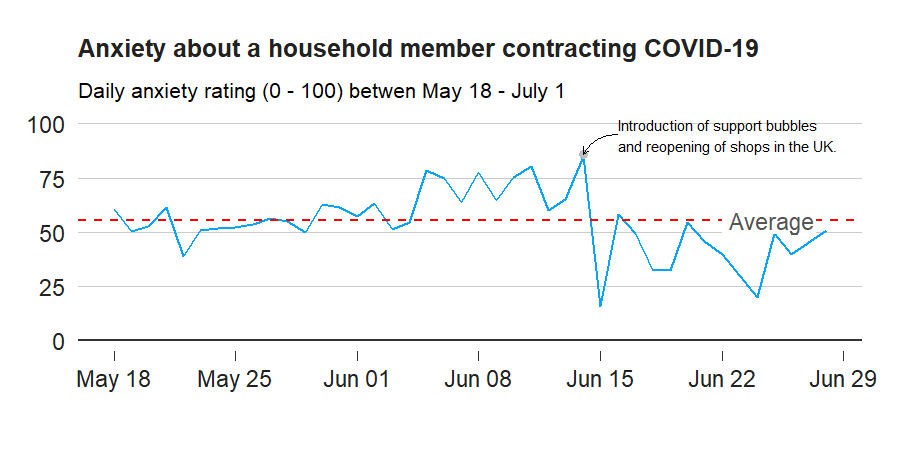

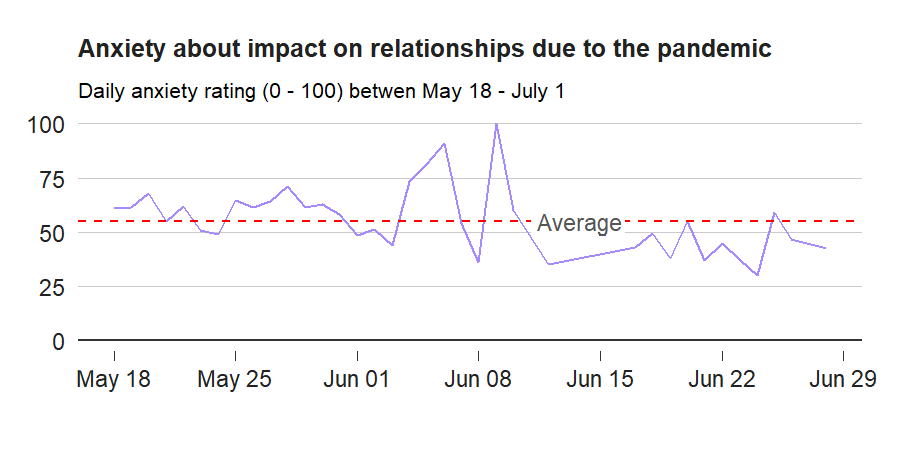

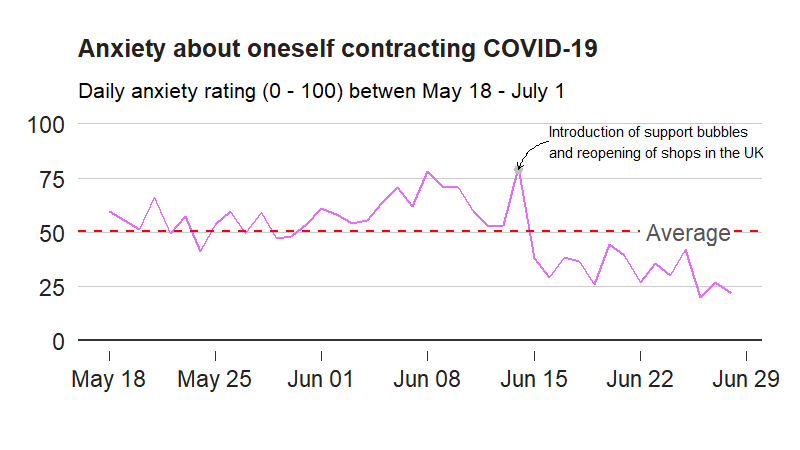

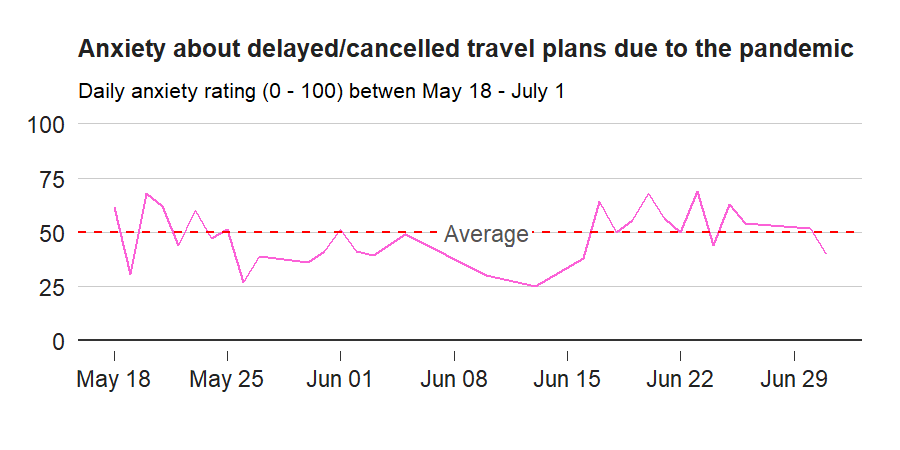

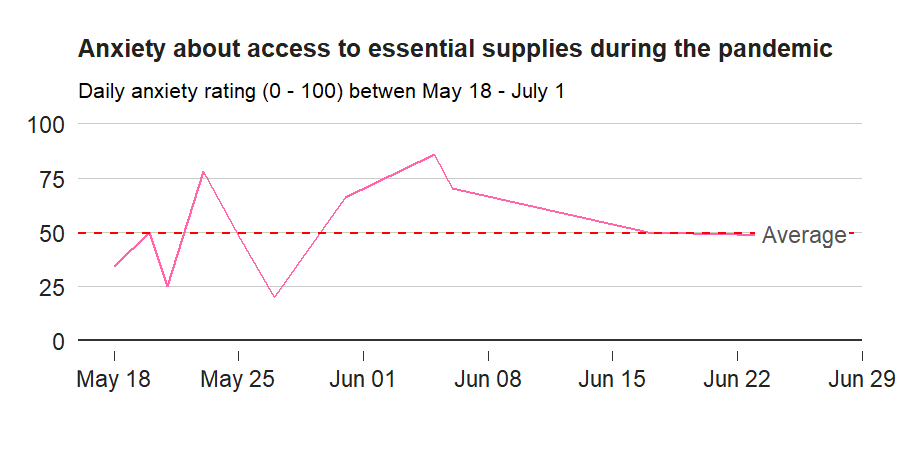
